# Proteome profiling of Cryoglobulinemia

**DOI:** 10.1101/2021.02.23.21252292

**Authors:** Peng Liu, Jianqiang Wu, Dandan Sun, Zhihong Qi, Xuzhen Qin, Wei Su

## Abstract

**Objective:** To better understand and identify candidate protein biomarkers of cryoglobulinemia (CGE) from healthy patients or benign brain disease control patients.

**Methods:** The tandem mass tag (TMT)-labeled quantitative proteomics approach was used to identify differential proteins in the serum proteome of 10 CGE patients and 10 disease control patients with negative cryoglobulin (CG), respectively. GO, KEGG and protein domain enrichment analysis was used for functional annotation of differential proteins. The biomarker candidates were validated by the parallel reaction monitoring (PRM) method in 20 samples (10 CGE patients, 5 healthy control patients, and 5 disease control patients).

**Results:** A total of 1004 proteins were quantitatively identified. Among these, 62 proteins were upregulated, and 56 proteins were downregulated. 25 candidate biomarkers were validated by PRM. 5 upregulated proteins of A2M, MMP2, VCAM1, MCAM, and PCSK9, and 4 downregulated proteins of C3, IGHG2, PROZ, PPBP were consistent with TMT results. Upregulated proteins mostly related with cholesterol metabolism, while downregulated proteins were associated with immune regulation.

**Conclusion:** This study is the first proteomic study of cryoglobulinemia in China These findings might contribute to the application of proteomics for detecting and monitoring CGE. And our findings indicated cholesterol metabolism accelerates the development of CGE.

---

Cryoglobulinemia (CGE) is a group of clinical syndromes caused by the presence of cryoglobulin in serum. Cryoglobulin (CG) is a special immunoglobulin that precipitates at low temperature and then dissolves at about 37°C. According to the different types and composition of CG, cryoglobulinemia is divided into three types, of which type I is mostly composed of monoclonal IgM, type II is composed of two different types of polyclonal IgG and monoclonal IgM, and type III is composed of the combination of polyclonal IgG and polyclonal IgM. According to different etiology, MC can be further divided into Essential Mixed Cryoglobulinemia (EMC) and secondary MC. The former has no clear etiology and accounts for about 10% of all MC patients. The latter is the main type of MC patients, mainly seen in infectious diseases such as Hepatitis C virus (HCV) and Hepatitis B virus, as well as autoimmune diseases such as systemic lupus erythematosus and primary Sjogren’s syndrome [1].

Previous studies have suggested that cryoglobulinemia is a rare disease (less than 5 in 10,000 people in Europe and North America). However, with the development of research, it has been gradually found that in some regions, such as the Mediterranean region, the incidence rate reaches 20%^[2]^, while 30-50% of HCV infected patients, 10% of systemic lupus erythematosus patients, and 5-20% of primary Sjogren’s syndrome patients all have MC^[1]^. Due to the relatively high prevalence of hepatitis C in China, according to China Statistical Yearbook, the annual incidence of hepatitis C is more than 200,000 in the past six years. Therefore, in theory, the prevalence of cryogulinemia in China is higher than that in other regions. However, it is easy to be ignored by clinicians due to the absence of any symptoms at the early stage or non-specific clinical manifestations (more than 60% present palpable purpura, arthralgia and fatigue) ^[3]^. As the disease progresses, severe irreversible clinical manifestations such as glomerulonephritis, peripheral neuropathy, and skin ulcers occur, and the 10-year survival rate was significantly reduced. At the same time, patients with MC have poor response to treatment, high mortality rate and sequelae rate of chronic organ failure (such as chronic renal failure and lymphoma) ^[4,5]^. Therefore, cryoglobulinemia is a disease with atypical symptoms and serious injury to patients. It is critical to find accurate early diagnosis and risk assessment markers for the prognosis of patients.

Mass spectrometry is a powerful tool to identify specific proteins from hundreds of protein species, allowing the study of biomarker screening in case-control research^[6,7]^. In the present study, we performed a quantitative proteomics study of CGE patients to investigate the possible candidate proteins for differential diagnosis.

## Methods

### Patients and Sample collection

A retrospective review of samples from ten cases of cryoglobulinemia (CGE) and matched 10 control samples from patients with negative cryoglobulin (CG) results in the same period was performed by the tandem mass tag (TMT)-labeled quantitative proteomics approach from Peking Union Medical College Hospital (PUMCH) of China. For validation purposes, an independent case-control test set (10 CG patients, 10 healthy controls) was obtained from the same hospital and tested by the parallel reaction monitoring (PRM) method.

A total of 1 mL of serum sample after clinical measurement was collected for each subject and stored at −80 °C. Cryoglobulin were tested by capillary electrophoresis typing method in PUMCH referring to previous reported methods^[8,9]^. In all cases, routine evaluation, including blood cell count, biochemical profile was reviewed. Clinical information was obtained from the Hospital Information System. The Institutional Ethnics Committee at the PUMCH approved the study (S-K904). An informed written consent was waived for remained samples of routine tests.

### Sample preparation

The cellular debris of serum sample was removed by centrifugation at 12,000 g at 4 °C for 10 min. Then, the supernatant was transferred to a new centrifuge tube. The top 12 high abundance proteins were removed by Pierce™ Top 12 Abundant Protein Depletion Spin Columns Kit (Thermo Fisher). Finally, the protein concentration was determined with BCA kit according to the manufacturer’s instructions. For digestion, the protein solution was reduced with 5 mM dithiothreitol for 30 min at 56 °C and alkylated with 11 mM iodoacetamide for 15 min at room temperature in darkness. The protein sample was then diluted by adding 100 mM TEAB to urea concentration less than 2M. Finally, trypsin was added at 1:50 trypsin-to-protein mass ratio for the first digestion overnight and 1:100 trypsin-to-protein mass ratio for a second 4 h-digestion.

### TMT-labeled quantitative proteomic analysis

After trypsin digestion, peptide was desalted by Strata X C18 SPE column (Phenomenex) and vacuum-dried. one unit of TMT/iTRAQ reagent were thawed and reconstituted in acetonitrile. The peptide mixtures were then incubated for 2 h at room temperature and pooled, desalted and dried by vacuum centrifugation. The tryptic peptides were fractionated into fractions by high pH reverse-phase HPLC using Thermo Betasil C18 column (5 μm particles, 10 mm ID, 250 mm length). Briefly, peptides were first separated with a gradient of 8% to 32% acetonitrile (pH 9.0) over 60 min into 60 fractions. Then, the peptides were combined into 6 fractions and dried by vacuum centrifuging.

### LC-MS/MS Analysis

The tryptic peptides were dissolved in 0.1% formic acid (solvent A), directly loaded onto a homemade reversed-phase analytical column (15-cm length, 75 μm i.d.). The gradient was comprised of an increase from 6% to 23% solvent B (0.1% formic acid in 98% acetonitrile) over 26 min, 23% to 35% in 8 min and climbing to 80% in 3 min then holding at 80% for the last 3 min, all at a constant flow rate of 400 nL/min on an EASY-nLC 1000 UPLC system.

The peptides were subjected to NSI source followed by tandem mass spectrometry (MS/MS) in Q ExactiveTM Plus (Thermo) coupled online to the UPLC. The electrospray voltage applied was 2.0 kV. The m/z scan range was 350 to 1800 for full scan, and intact peptides were detected in the Orbitrap at a resolution of 70,000. Peptides were then selected for MS/MS using NCE setting as 28 and the fragments were detected in the Orbitrap at a resolution of 17,500. A data-dependent procedure that alternated between one MS scan followed by 20 MS/MS scans with 15.0s dynamic exclusion. Automatic gain control (AGC) was set at 5E4. Fixed first mass was set as 100 m/z.

### Database Search

The resulting MS/MS data were processed using Maxquant search engine (v.1.5.2.8). Tandem mass spectra were searched against human uniprot database (www.uniprot.org) concatenated with reverse decoy database. Trypsin/P was specified as cleavage enzyme allowing up to 4 missing cleavages. The mass tolerance for precursor ions was set as 20 ppm in First search and 5 ppm in Main search, and the mass tolerance for fragment ions was set as 0.02 Da. Carbamidomethyl on Cys was specified as fixed modification and acetylation modification and oxidation on Met were specified as variable modifications. FDR was adjusted to < 1% and minimum score for modified peptides was set > 40.

After filtering, peptide abundances in the different reporter ion channels were normalized. The protein abundance ratio was based on unique peptide results. For differential protein filtering, proteins with high inter-individual variations within each group (top 5% inter-individual CV) were excluded. The relative quantification values of each sample made log2 changes to normalize the data, then corresponding p-values were tested. When p-value<0.05, differentially expressed value of proteins over 1.3 is set as the upregulated cutoff, while, less than 1/1.3 is set as the downregulated cutoff.

### Bioinformatics Methods

Proteins were classified by Gene Ontology annotation based on three categories: biological process, cellular component and molecular function. For further hierarchical clustering based on differentially expressed protein functional classification (including: GO, Domain, Pathway, Complex). All differentially expressed protein database accession or sequence were searched against the STRING database version 10.1 for protein-protein interactions.

### Parallel reaction monitoring (PRM) analysis

For PRM validation, 25 distinct differential proteins from LC-MS/MS analysis were evaluated using 20 validation group samples. Each sample was analyzed in schedule mode. To ensure the quality of the data, the mixed sample was analyzed as quality control to ensure the stability of the instrument signal during the whole process. The iRT standard peptide analysis was also added to each sample, and the stability of chromatographic retention time was assessed during the analysis. Two technical repeats were run in each sample. To reduce system bias, different groups of the samples were analyzed in a random order for mass spectrometry analysis.

In PRM analysis, proteins were extracted and digested as described above. The gradient was comprised of an increase from 6% to 23% solvent B (0.1% formic acid in 98% acetonitrile) over 38 min, 23% to 35% in 14 min and climbing to 80% in 4 min then holding at 80% for the last 4 min, all at a constant flow rate of 700 nL/min on an EASY-nLC 1000 UPLC system. The peptides were subjected to NSI source followed by tandem mass spectrometry (MS/MS) in Q ExactiveTM Plus (Thermo) coupled online to the UPLC. The electrospray voltage applied was 2.0 kV. The m/z scan range was 350 to 1000 for full scan, and intact peptides were detected in the Orbitrap at a resolution of 35,000. Peptides were then selected for MS/MS using NCE setting as 27 and the fragments were detected in the Orbitrap at a resolution of 17,500. A data-independent procedure that alternated between one MS scan followed by 20 MS/MS scans. Automatic gain control (AGC) was set at 3E6 for full MS and 1E5 for MS/MS. The maximum IT was set at 20 ms for full MS and auto for MS/MS. The isolation window for MS/MS was set at 2.0 m/z.

The resulting MS data were processed using Skyline (v.3.6). Peptide settings: enzyme was set as Trypsin [KR/P], Max missed cleavage set as 2. The peptide length was set as 8-25, Variable modification was set as Carbamidomethyl on Cys and oxidation on Met, and max variable modifications was set as 3. Transition settings: precursor charges were set as 2, 3, ion charges were set as 1, 2, ion types were set as b, y, p. The product ions were set as from ion 3 to last ion, the ion match tolerance was set as 0.02 Da. All data were imported into the Skyline software, and the correct peaks were selected manually, and all the peptide results in all samples were exported. Total ionic chromatography (TIC) of ions with +2 − +5 charge was extracted from each sample using Progenesis software. The mass spectra of each peptide in each sample were normalized with the TIC strength of the sample to adjust the errors from the sample loading amount and MS signal intensity. The results of each peptide were quantitatively analyzed, and the differential proteins between different groups were analyzed and compared with TMT results.

### Statistics

Pattern recognition analysis (principal component analysis, PCA; orthogonal partial least squares discriminant analysis, OPLS-DA) was performed using SIMCA 14.0 (Umetrics, Sweden) software. Comparison of the selected peptides between groups was analyzed using the paired t-test when appropriate. P < 0.05 was considered significant.

## Results

### Demographic of included patients

A total of 20 cryoglobulinemia patients were included in this study, 10 in TMT analysis and 10 in RPM analysis. To screen the accurate target protein, 15 gender and age matched disease controls with negative CG results and 5 healthy control were respectively included in TMT and RPM analysis. The main disease in the disease control group covered purpura, nephritis, et ct. Hepatitis C antibody and HIV antibody were all negative.While, in CGE patients and disease control group there were 20-40% patients with positive Hepatitis B antigen results (shown in Supplemental Table 1).

**Table 1.** Demographic of included patients in TMT analysis.

| Analysis | Group | N | Age | Male/Gender | CG quantity |
| --- | --- | --- | --- | --- | --- |
| TMT | Cryoglobulinemia | 10 | 58.5±9.4 | 5/10 | 3.38% |
|  | Disease Control | 10 | 56.9±7.1 | 4/10 | - |
| PRM | Cryoglobulinemia | 10 | 49.5±10.5 | 5/10 | 2.33% |
|  | Disease Control | 5 | 50±16.1 | 2/5 | - |
|  | Healthy Control | 5 | 54±8.6 | 3/5 | - |

### Differential proteins identified by TMT-based proteomics

The general experimental workflow is illustrated in Fig. 1. A total of 11154 peptides and 1004 proteins were quantitatively identified in samples from the CGE and disease control groups, of which 10768 peptides were unique peptides and 920 proteins could be quantified. Compared with the control group, 62 proteins were upregulated, and 56 proteins were downregulated. Principal component analysis (PCA), relative standard deviation (RSD) and Pearson’s Correlation Coefficient were used to evaluate the quantitative repeatability of proteins. Since three types of CGE patients were included in the CGE group, PC of CGE group was higher than PC of control group (23.4% explained var. vs. 15.5% explained var.), and PCA analysis indicated there were overlapping between these two groups. RSD distribution box figure of two groups were shown in Figure 1A. The median inter-individual CV of CEG group and disease control group was separately 22.9%, and 19.5%, indicating low variation levels within the groups.

**Figure 1:**
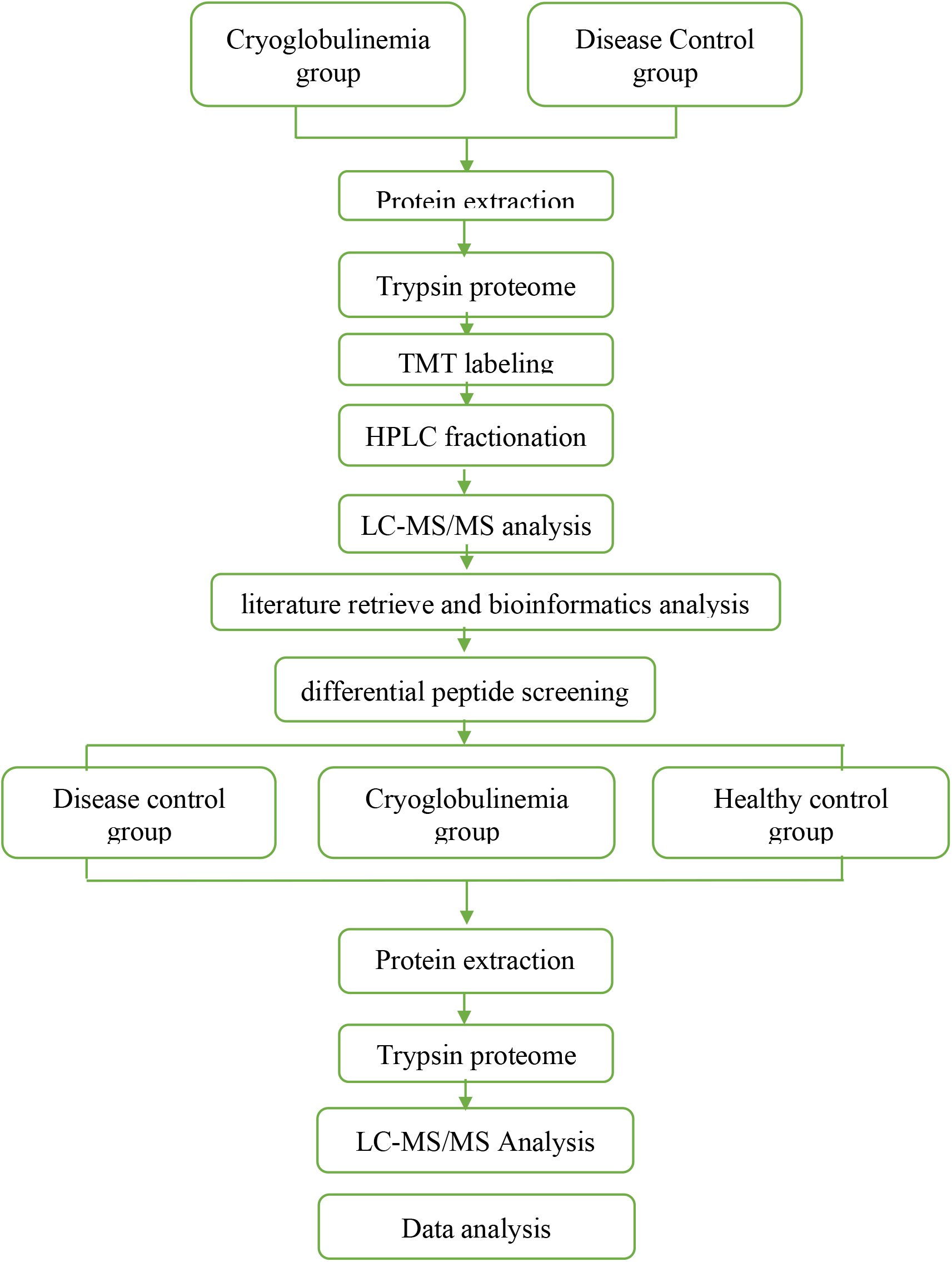
Workflow of this study

To elucidate functional differences between the upregulated and downregulated proteins, the quantified proteins were analyzed by Gene ontology (GO) analysis. These proteins were mainly involved in cellular process and single-organism process, and have binding and catalytic molecular function. 54.24% of the differentially expressed proteins were located in extracellular (Supplemental Fig. 1).

The quantified proteins were analyzed by three types of enrichment-based clustering analyses to find the significant enrichment tendency in some functions: Gene ontology (GO) enrichment-based clustering analysis, protein domain enrichment-based clustering analysis and KEGG pathway enrichment-based clustering analysis. In the biological process, the proteins associated with protein-lipid complex assembly, regulation of cellular ketone metabolic process, and lipid transport were enriched in the differentially expressed proteins (Figure 2B). KEGG pathway analysis revealed that NF-kappa B signaling pathway, African trypanosomiasis pathway, Cholesterol metabolism pathway were significantly enriched in the differentially expressed proteins (Figure 2C). Protein domains of the differentially expressed proteins mainly presented in immunoglobulin-like domain and immunoglobulin-like fold (Figure 2D).

**Figure 2:**
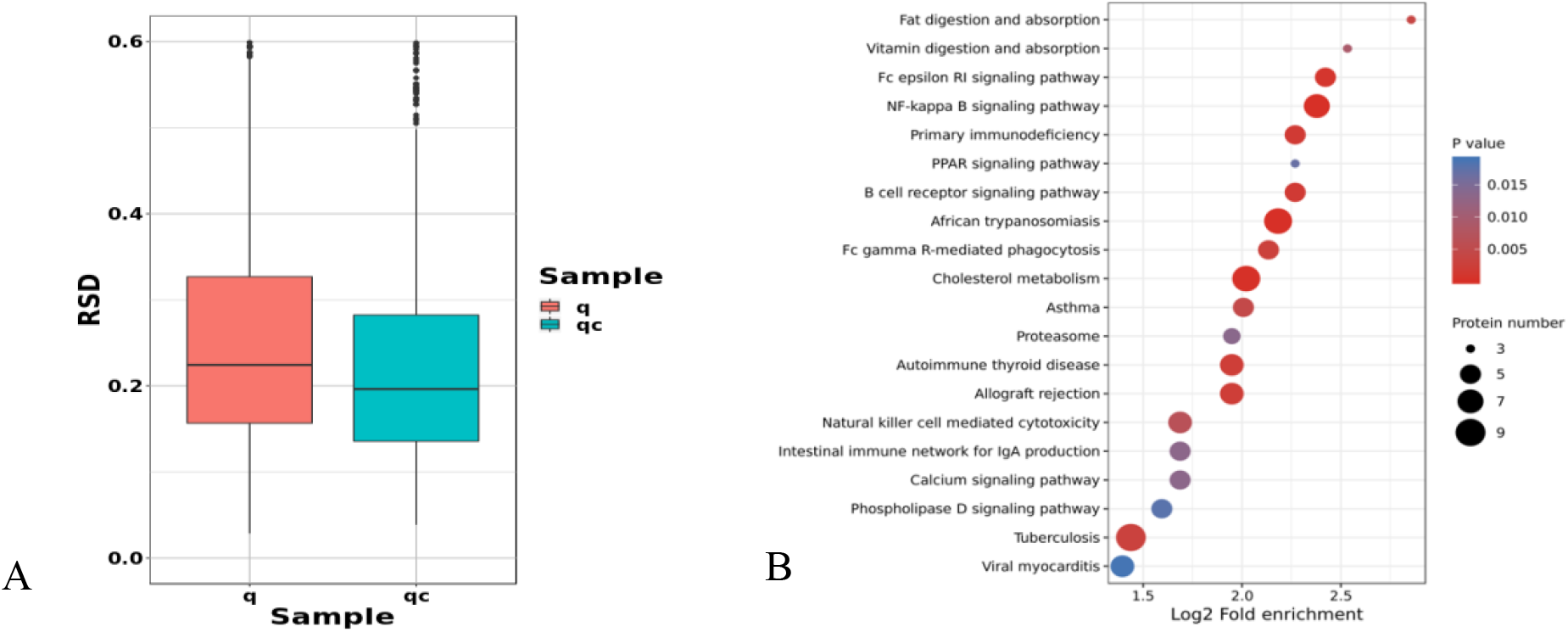
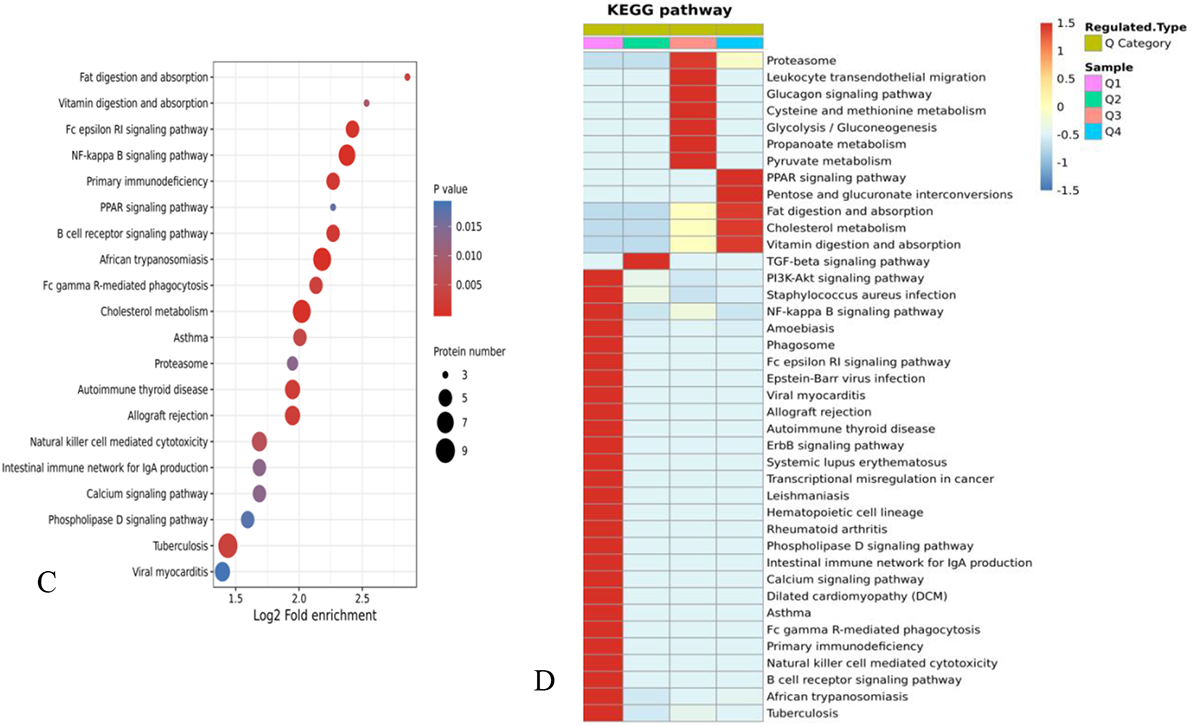
TMT-based proteomic analysis of MB. A: RSD distribution box figure of repeated samples. q: CGE group, qc: disease control group. B: The results of the top 20 classifications with the most significant enrichment are given in the bubble chart. In the bubble diagram, the vertical axis is functional classification or pathway, and the horizontal axis is Log2 converted value of the ratio of the difference protein in the functional type to the change of the ratio of the identified protein. The color of the circle represents the p-value of enrichment significance, and the size of the circle represents the number of differential proteins in functional classes or pathways. C: The differentially expressed proteins were enriched and distributed in bubbles in KEGG pathway. D: Heat map of cluster analysis based on KEGG pathway enrichment.

All differentially expressed proteins were divided into four quantiles (Q1-Q4) according to differential expression multiples as follows: Q1 (≤ 0.667), Q2 (0.667 < ratio ≤ 0.769), Q3 (1.3 < ratio ≤ 1.5) and Q4 (>1.5) to determine the association with various differential expression multiples proteins. Red indicates strong enrichment, and blue indicates weak enrichment. Proteins in PPAR signaling pathway, pentose and glucuronate interconversions, fat digestion and absorption, cholesterol metabolism and vitamin digestion and absorption were instinctively enriched and expressed higher in CGE group than in disease control group. Based on these results, 25 candidate proteins with over 3 unique peptides identified and high differential expression multiples in the PPAR signaling pathway, pentose and glucuronate interconversions, fat digestion and absorption, cholesterol metabolism and vitamin digestion and absorption pathways were selected for further validation by PRM analysis in independent samples.

### Validation of differentially expressed proteins by PRM-based proteomics

22 of 25 proteins were identified by PRM-based proteomics. 5 upregulated proteins of A2M, MMP2, VCAM1, MCAM, and PCSK9, and 4 downregulated proteins of C3, IGHG2, PROZ, PPBP were consistent with TMT results, no matter whether control group contains disease control or healthy control. Upregulated proteins mostly related with cholesterol metabolism, while downregulated proteins were associated with immune regulation.

**Table 2.** Comparison between TMT and PRM results.

| Protein Accession | Protein Gene | Protein description | CGE/disease control and healthy control Ratio (PRM) | CGE/disease control Ratio (PRM) | CGE/disease control Ratio(TMT) |
| --- | --- | --- | --- | --- | --- |
| P00739 | HPR | Haptoglobin-related protein | 0.79 | 1.01 | 0.49 |
| P01023 | A2M | Alpha-2-macroglobulin | 2.05 | 2.20 | 1.36 |
| P01024 | C3 | Complement C3 | 0.86 | 0.96 | 0.80 |
| P01859 | IGHG2 | Immunoglobulin heavy constant gamma 2 | 0.75 | 0.79 | 0.62 |
| P02654 | APOC1 | Apolipoprotein C-I | 0.84 | 1.17 | 2.07 |
| P02655 | APOC2 | Apolipoprotein C-II | 0.82 | 1.02 | 1.62 |
| P02656 | APOC3 | Apolipoprotein C-III | 1.15 | 0.93 | 1.60 |
| P02743 | APCS | Serum amyloid P-component | 0.85 | 1.12 | 0.66 |
| P08253 | MMP2 | 72 kDa type IV collagenase | 1.22 | 1.35 | 1.46 |
| P08519 | LPA | Apolipoprotein(a) | 2.00 | 0.99 | 1.35 |
| P0DOY3 | IGLC3 | Immunoglobulin lambda constant 3 | 1.16 | 0.93 | 0.50 |
| P18428 | LBP | Lipopolysaccharide-binding protein | 0.84 | 0.53 | 1.41 |
| P19320 | **VCAM1** | Vascular cell adhesion protein 1 | 2.12 | 1.82 | 1.38 |
| P22891 | **PROZ** | Vitamin K-dependent protein Z | 0.61 | 0.90 | 0.71 |
| P22897 | MRC1 | Macrophage mannose receptor 1 | 1.97 | 0.82 | 1.47 |
| P43121 | **MCAM** | Cell surface glycoprotein MUC18 | 1.26 | 1.32 | 1.32 |
| P49913 | CAMP | Cathelicidin antimicrobial peptide | 0.60 | 0.92 | 1.51 |
| P55058 | PLTP | Phospholipid transfer protein | 0.90 | 0.97 | 1.22 |
| Q8NBP7 | **PCSK9** | Proprotein convertase subtilisin/kexin type 9 | 1.04 | 1.17 | 1.22 |
| P02775 | **PPBP** | Platelet basic protein | 0.58 | 0.64 | 0.77 |
| P55103 | INHBC | Inhibin beta C chain | 0.94 | 0.82 | 1.20 |
| Q9NPH3 | IL1RAP | Interleukin-1 receptor accessory protein | 0.97 | 1.26 | 1.06 |

The bold font indicates that the expression ratio trend is consistent in different control groups

## Discussion

In this study, we enrolled 20 CGE patients, 15 disease controls and 5 healthy controls to investigate the differential proteins between three groups with TMT-based proteomics and PRM-based proteomics. A total of 1004 proteins were quantitatively identified. Among these, 62 proteins were upregulated, and 56 proteins were downregulated. These proteins were mainly involved in cellular process and single-organism process, and have binding and catalytic molecular function. In the biological process, the proteins associated with protein-lipid complex assembly, regulation of cellular ketone metabolic process, and lipid transport were enriched in the differentially expressed proteins. KEGG pathway analysis revealed that NF-kappa B signaling pathway, African trypanosomiasis pathway, Cholesterol metabolism pathway were significantly enriched in the differentially expressed proteins. 22 of 25 proteins were identified by PRM-based proteomics. 5 upregulated proteins of A2M, MMP2, VCAM1, MCAM, and PCSK9, and 4 downregulated proteins of C3, IGHG2, PROZ, PPBP were consistent with TMT results, no matter whether control group contains disease control or healthy control. Upregulated proteins mostly related with cholesterol metabolism, while downregulated proteins were associated with immune regulation.

Since cryoglobulinemia is a rare disease, few studies of proteomics have been conducted in this field. Ogishi et al. isolated Fab fragments from resolubilized cryoprecipitate and serum remnant and labeled the Fab fragments with the isobaric tag for relative and absolute quantification (iTRAQ) for quantitative comparison. They identified that immunoglobulin variable heavy chain (VH) sequences highly enriched in pathological immune complex from clinical specimens obtained from a patient with hepatitis C virus-induced cryoglobulinemia^[10]^. While, in our study we found immunoglobulin related proteins were mostly downregulated in CGE group compared with control group. The unique patient with HCV infection in the previous study was obviously different with this study, resulting in the inconsistency of research results. Lee et al. reported a novel bedside-to-bench proteomic approach through heavy-chain sequencing protocol to type cryoglobulins in term of their unique molecular barcodes^[11]^. This is a good application of proteomics for CGE diagnosis and treatment monitoring. Using paraffin-embedded tissue, cryoglobulinemic glomerulonephritis (GN) were compared with amyloidosis, fibrillary GN, and immunotactoid glomerulopathy. The absence of apolipo-protein E correlates with the lack of fibrillar deposits in cryoglobulinemic GN. We found Apo CI, Apo CII, Apo CIII and Apo E were increased in CGE compared with disease control without the presence of Apo E^[12]^. Mass spectrometry-based proteomic analyzed cutaneous cryoglobulinemia in formalin-fixed tissues. They found thrombi contained immunoglobulins and the results were consistent with the prior clinical diagnosis and laboratory data^[13]^.

Previous studies were mostly focused on immunoglobulins related proteins for the application of proteomics. While in our study we noted obvious upregulated proteins involved in cholesterol metastasis. Few studies have been conducted on the pathogenesis of CGE with the relationship of cholesterol metastasis, but its role in lymphatic proliferative diseases has been reported. Proteins involved in the formation and transport of free fatty acids (FFA), triglyceride (TG) and total cholesterol (TC) were significantly increased in the peripheral blood of chronic lymphocytic leukemia (CLL) patients. For example, fatty acid binding protein 3 (FABP3) in the PPARα signaling pathway, are overexpressed in CLL patients, which proves that CLL patients can absorb exogenous lipids from the extracellular through CD36 receptor^[14]^. In lymphoproliferative diseases, it has been shown that there is a correlation between abnormal lipid metabolism and lymphocyte proliferation. Epidemiological study of patients with Chronic lymphocyte leukemia (CLL) found that 17.6% patients with CLL had abnormal lipid metabolism. Multivariate analysis showed that abnormal lipid metabolism was related to CLL (p < 0.001). lipid-lowering treatment is helpful for the survival of high CLL^[15]^. Compared with children with Acute B-lymphoblastic leukemia (B-ALL), obese children are more likely to relapse and have a poorer prognosis. Patients with obesity, type 2 diabetes, hyperglycemia and other risk factors, the risk of progression to B cell malignant disease is significantly increased. In vitro studies have shown that if nutrient supply is restricted, it can prevent the malignant transformation of precursor B cells. In the B-ALL rat model, hunger can slow down the progression of disease^[16]^. The possible reason may be that the process of B lymphocyte activation and antibody production requires metabolic regulation to meet the demand for increased energy. Studies have shown that during activation, B cells can counterbalance a temporary increase in energy expenditure, thereby ensuring immune homeostasis. If autoreactive B cell receptors are permanently overactivated, it will develop metabolic abnormalities and pathological B cell proliferation^[17]^.

The limitation of our study is the limited verification samples due to the expensive testing, and the healthy controls are not matched with the number of disease. So, the selection bias may influence the strength of our study. However, the sample size is larger than previous studies and our study is the first proteomic study of CGE in China. Therefore, it is instructive for CGE pathogenesis research and biomarker screening.

## Data Availability

To access all data referred to in the manuscript should get approval of the corresponding author

## Acknowledgements

The manufacturer provided equipment and reagents for this study but had no role in directing the study or influencing the study outcomes.

## Declarations

We declare that the authors have no competing interests, or other interests that might be perceived to influence the results and/ or discussion reported in this paper. The results/data/figures in this manuscript have not been published elsewhere, nor are they under consideration. All materials are owned by the authors and/or no permissions are required. We are responsible for the submission of this article and accept the conditions of submission and the BMC Copyright and License Agreement.

## Authors’ contributions

PL proposed this study; JQ W helped the proteomic tests; DD S collected the samples; XZ Q wrote the draft; WS conducted the electrophoresis tests and contacted with clinicians. All authors have accepted responsibility for the entire content of this manuscript and approved its submission.

## Ethics approval and consent to participate

This study was approved by the ethics committee of Peking Union Medical College Hospital (S-K904).

## Consent for publication

Informed consent was waived for the use in this study of the remaining samples left after diagnosis.

